# Cumulative hippocampal seizure-related burden impairs long-term memory consolidation in focal epilepsy

**DOI:** 10.64898/2026.05.20.26353420

**Authors:** Ionut-Flavius Bratu, Isabelle Lambert, Olivier Felician, Samuel Medina Villalon, Agnès Trébuchon, Fabrice Bartolomei

**Affiliations:** Department of Epileptology and Cerebral Rhythmology, Timone Hospital, 13005, Marseille, France; Systems Neuroscience Institute, Aix-Marseille University, French National Institute of Health and Medical Research (UMR 1106), 13005, Marseille, France; Department of Neurology and Neuropsychology, Timone Hospital, 13005, Marseille, France

**Author notes:** **Correspondence to:** Ionut-Flavius Bratu, Department of Epileptology and Cerebral Rhythmology, Timone Hospital, 264 Saint Pierre, 13005, Marseille, France.

## Abstract

**Objective:** Memory impairment is a frequent comorbidity of focal epilepsy, incompletely explained by seizure frequency or structural pathology. Ictal and postictal hippocampal dysfunction disrupt memory processes, but their cumulative impact remains poorly quantified. This study introduces cumulative hippocampal seizure-related burden metrics and examines their association with long-term memory consolidation.

**Methods:** Twenty consecutive patients undergoing stereo-EEG in Marseille (2016–2018) were prospectively included. Continuous stereo-EEG recordings between two memory assessments (30 minutes and one week post-encoding) were analysed. Hippocampal ictal involvement and durations were assessed using epileptogenicity markers and visual stereo-EEG analysis. The postictal period was quantified using permutation entropy. Cumulative hippocampal seizure-related burden metrics (ictal, postictal and combined: c-HipSZB) were computed across hippocampus-involving ictal events. Verbal and visual memory were assessed using standardized recall and recognition tasks. Associations were examined using univariate and multivariate analyses.

**Results:** Higher dominant-hemisphere hippocampal burden was associated with poorer one-week verbal memory (performance and retention), independently of most covariates. Higher c-HipSZB was associated with lower total recall performance (RT; free + cued) and RT retention (β = −25.04 and −23.88; R² = 0.57 and 0.53; p < 0.05) and accounted for the greatest variance in both outcomes (adjusted R² = 0.59 and 0.53; β = −25.45 and −24.27; p < 0.01), particularly when adjusting for epilepsy duration. No robust associations were observed between non-dominant-hemisphere hippocampal seizure-related burden metrics and visual memory. Effects predominantly involved recall.

**Interpretation:** Cumulative ictal–postictal hippocampal dysfunction is a major determinant of impaired long-term verbal memory consolidation in focal epilepsy.

## Introduction

Epilepsy is associated with frequent cognitive comorbidities spanning language, attention and memory.^1^ Among focal epilepsies, TLE is particularly associated with episodic memory impairment.^2^ Dominant-hemisphere TLE typically impairs verbal memory, while non-dominant-hemisphere TLE has a less consistent association with visual memory deficits.^3–8^ Because the hippocampus is central to memory, with memory traces encoded in the hippocampus and gradually consolidated in neocortical regions^9^, hippocampal dysfunction is considered a key contributor to memory impairment in TLE.^10^ Moreover, people with TLE frequently report disabling memory difficulties despite normal performance on standard neuropsychological tests.^11^ This discrepancy has been attributed to accelerated long-term forgetting, in which newly acquired information is initially retained, but abnormally decays over days to weeks, reflecting disrupted long-term consolidation.^12^ Although seizure frequency, epilepsy duration, hippocampal sclerosis^13^, interictal discharges^14^ and hippocampal ictal involvement^15^ have all been linked to cognitive decline, they do not fully explain interindividual variability in memory outcomes.^16^

Mechanistically, both ictal and postictal hippocampal dysfunction may impair memory consolidation.^10^ Seizures can disrupt hippocampal network activity causing transient amnesia for peri-ictal events.^17,18^ Moreover, during the postictal state – the period of abnormal brain activity following a seizure until return to baseline^19^ – patients can experience memory impairment.^20^ Experimental work showed that ictally triggered hippocampal vasoconstriction results in severe hypoxia-induced inhibition of long-term potentiation, a process essential for memory consolidation.^21^ This inhibition parallels postictal memory impairment, both potentially persisting for tens of minutes to several hours postictally. Despite these mechanistic insights, most clinical studies rely on coarse metrics (e.g., seizure count^14^), which may underestimate the cumulative functional impact of seizures on memory networks. To address this limitation, we introduce the concept of cumulative hippocampal seizure-related burden, defined as the total duration of ictal and postictal hippocampal dysfunction **(Fig. 1)**.

**Figure 1.**
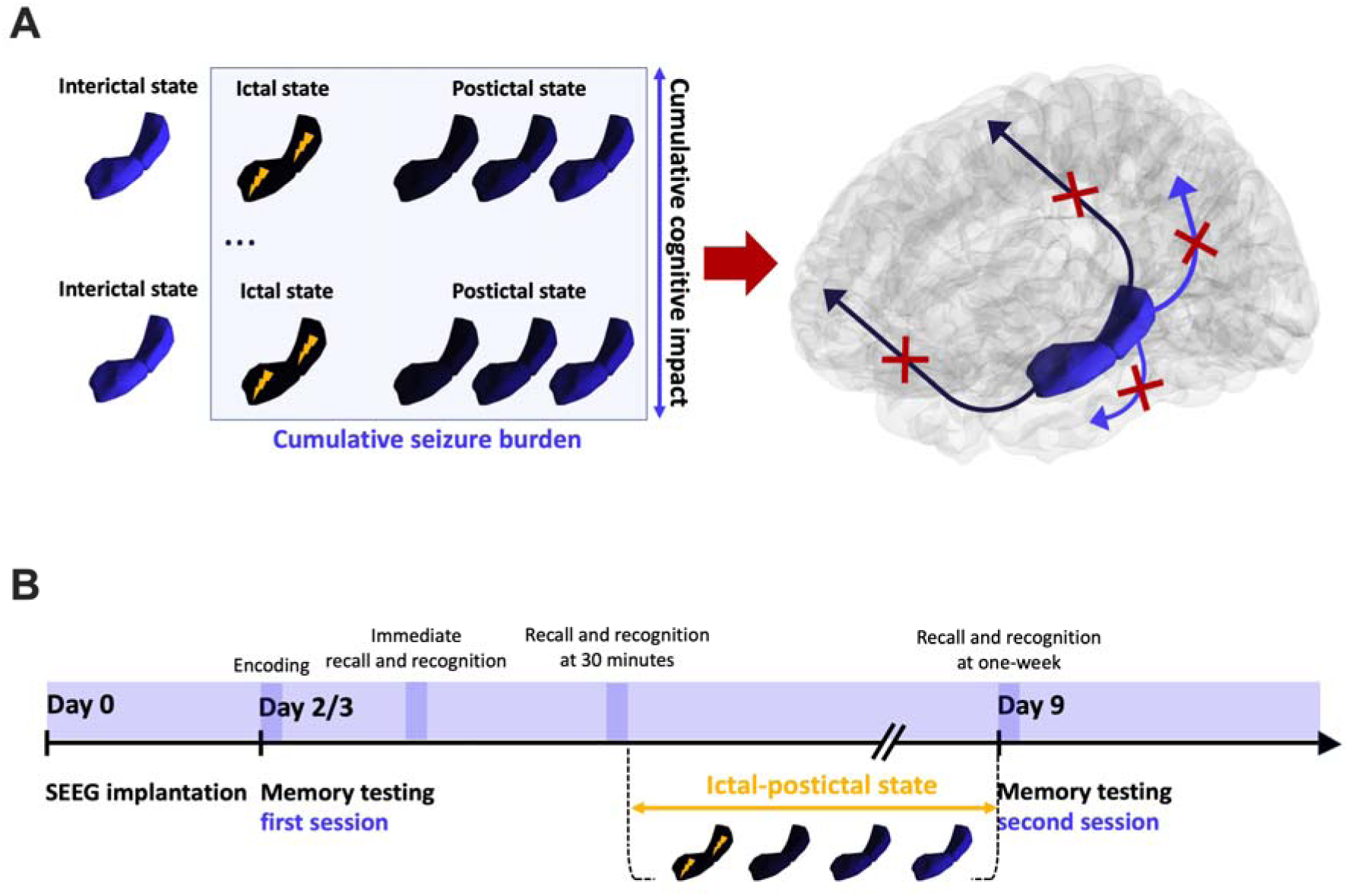
Conceptual framework and experimental design linking cumulative hippocampal seizure-related burden to long-term memory consolidation. **(A)** Repeated transitions of the hippocampus through ictal (lightning symbols) and postictal states over time contribute to an increasing cumulative seizure-related burden (x-axis), which parallels a progressive cumulative cognitive impact (y-axis). Recurrent hippocampal dysfunction during ictal and postictal states may impair memory consolidation by disrupting hippocampus–neocortical communication (arrows), ultimately weakening long-term memory trace formation (red crosses). **(B)** Schematic representation of the memory testing protocol and the integration of quantification metrics related to cumulative seizure-related burden in the hippocampus. Memory encoding was followed by immediate and 30-minute recall and recognition during the first testing session and by delayed recall and recognition at one week, allowing assessment of long-term memory consolidation. Ictal activity occurring between the two testing sessions contributes to the cumulative hippocampal seizure-related burden.

Quantifying such cumulative metrics requires approaches capable of capturing the full spectrum of peri-ictal hippocampal activity dynamics. Given the nonlinearity of brain electrical activity as reflected in stereo-electroencephalography (SEEG) signals, permutation entropy (PE) provides a signal complexity measure that is sensitive to these dynamics.^22^ PE-based analyses have previously enabled SEEG quantification of the postictal period^19^ and a deeper understanding of the mechanisms underlying postictal deficits.^23^

Lambert et al.^14^ demonstrated that hippocampal interictal activity during non-rapid eye movement (NREM) sleep and seizure count are associated with long-term forgetting in TLE. Extending this work, we hypothesized that cumulative seizure-related hippocampal dysfunction is a major determinant of long-term consolidation deficits in focal epilepsy. To test this, ictal and postictal burden metrics were derived from hippocampus-involving SEEG-recorded ictal events between two memory testing sessions (at 30 minutes and one week after encoding). Memory was assessed using both verbal and visual tasks, including recall- and recognition-based measures, allowing evaluation of long-term performance and retention. Accounting for relevant clinical covariates, we examined the association between hippocampal seizure-related burden metrics and memory outcomes within a hypothesis-driven framework—linking dominant-hemisphere hippocampal involvement to verbal memory and non-dominant-hemisphere involvement to visual memory.

## Materials and methods

### Patient selection

Patients undergoing SEEG evaluation at Timone Hospital, Marseille (June 2016–May 2018) were prospectively recruited if they had hippocampal SEEG sampling and were able to participate in memory assessment (Lambert et al.^14^).

### Presurgical evaluation

Non-invasive investigations included medical history, clinical examination, neuropsychological assessment, cerebral structural magnetic resonance imaging (MRI) and 18-fluorodeoxyglucose-positron emission computed tomography (PET-CT), as well as prolonged video-EEG monitoring. Additional investigations (e.g., language functional MRI) were performed case-by-case. The study complied with the Declaration of Helsinki. All participants provided written informed consent and the protocol was approved by the French Institute of Health (IRB-15226).

Invasive SEEG evaluations followed an electro-clinical, hypothesis-driven approach, in accordance with national guidelines^24^. Implantations were carried out using 10–18-contact intracerebral electrodes (Alcis, France; contact length: 2 mm; intercontact spacing: 1.5 mm; diameter: 0.8 mm). Post-implantation cerebral CT and/or MRI scans were systematically obtained to verify electrode positioning and detect periprocedural complications. SEEG signals were recorded using 128- or 256-channel acquisition systems (Deltamed Natus Medical), with a high-pass filter at 0.16 Hz (−3 dB) and an anti-aliasing low-pass filter at 97/170/340/680 Hz (corresponding to sampling rates of 256/512/1024/2048 Hz).

### SEEG contact localisation

Pre-SEEG cerebral T1-weighted MRI scans were processed for volumetric segmentation and cortical surface reconstruction using FreeSurfer.^25^ Anatomical parcellation was performed using the Virtual Epileptic Patient (VEP) atlas (https://ins-amu.fr/vep-atlas).^26^ Co-registration of cerebral scans (pre-implantation MRI and post-implantation CT) employed GARDEL software^27^ (https://meg.univ-amu.fr/doku.php?id=epitools:gardel), enabling semi-automatic detection and anatomical localisation of SEEG contacts by projection into the patient’s own cerebral MRI space and assignment to VEP atlas parcels^25^.

### Memory assessment

A two-session experimental protocol was designed to assess long-term memory consolidation^14^ using visual and verbal tasks. The first session was conducted 48–72 hours after SEEG implantation, depending on the patient’s clinical condition and included encoding, as well as immediate and 30-minute delayed recall and recognition. For all tests, encoding was reinforced until ≥80% correct responses were achieved, with a maximum of three attempts. The second session took place seven days later and assessed recall and recognition. Participants were not informed of this delayed assessment to prevent rehearsal of the memorised information between sessions. Memory testing at a 24-hour delay was not evaluated to prevent rehearsal effects induced by repeated testing, which could artificially reduce forgetting at the one-week delay. Assessments were performed by two neuropsychologists and one neurologist experienced in psychometric testing (IL).

#### Verbal memory

Verbal memory was evaluated using the RL/RI-16 (French Free and Cued Selective Reminding Test)^28^ and the Logical Memory subtest of the Wechsler Memory Scale—Third Edition (WMS-III)^29^. The RL/RI-16^29^ consists of a 16-word list, with each word belonging to a distinct semantic category. During encoding, words were presented on a screen divided into four quadrants, each containing one word. For each word, a unique semantic category cue was provided orally and participants identified, pointed to and read aloud the corresponding word. After correct identification of the four words of a given set, the visual support was removed and an immediate cued recall was performed, during which the semantic cues were presented again and participants were asked to recall the corresponding words. In case of error, words were re-presented with their cue. Each four-word set could be repeated up to three times to ensure adequate encoding before proceeding to the next set, in accordance with the RL/RI-16 procedure and the study encoding protocol. After a 20-second backward counting task used to limit rehearsal, retrieval was assessed using free recall followed by semantic cued recall for non-retrieved items. This procedure was repeated across three trials, each separated by backward counting. The total recall score at the third trial was considered as the immediate recall measure. Total recall (RT), reflecting overall retrieval performance after controlled encoding, was calculated as the sum of free (RL) and cued (RI) recall

Verbal memory was additionally assessed using the Logical Memory subtest of the WMS-III^28^ (WMS-III-LM). Two short narrative stories were read aloud according to standardised procedures. Participants were asked to recall each story immediately after presentation and again after a 30-minute delay. Performance was scored based on correctly reproduced story units (25 per story). Following delayed recall, a recognition task was administered consisting of 15 two-alternative forced-choice questions probing specific narrative elements (e.g. characters, actions or contextual details).

#### Visual memory

Visual memory was assessed using a modified Delayed Matching-to-Sample task (DMS-48)^30^ and the Visual Reproduction (Abstract Drawings) subtest of the WMS-III (WMS-III-VR)^28^. During encoding, participants were sequentially presented with 48 visual stimuli (targets), each displayed in one of four predefined screen quadrants, with spatial positions balanced across trials. To ensure perceptual processing, participants performed a simple judgement for each image (biological versus human-made) and indicated its quadrant. After an interference interval, recognition was assessed using 96 single-item trials (48 targets and 48 novel distractors) presented in random order. For each image, participants indicated whether they had seen it before. For recognised items, they indicated the screen quadrant of presentation during the encoding phase. Performance was quantified as recognition accuracy and, for correctly recognised items, spatial position accuracy, providing measures of visual recognition and contextual memory.

Visual memory was also assessed using the Visual Reproduction subtest of the WMS-III^28^. During encoding, drawings were presented for 10 seconds and participants were asked to reproduce them. The task followed standard procedures, with the addition of encoding reinforcement identical to that applied in the other memory tests. 30 minutes after encoding, following completion of a non-visual task, participants were asked to reproduce the learned drawings. A recognition task was then administered, in which seven target and 14 distractor drawings were presented and participants indicated whether each had been previously seen.

### SEEG signal analysis

All signal analyses were performed in AnyWave software^31^ (https://meg.univ-amu.fr/AnyWave/download.html) in bipolar montage.

Hippocampal ictal involvement was assessed using a combined visual and quantitative SEEG approach based on the Epileptogenicity Index (EI)^32^ and the Connectivity Epileptogenicity Index (cEI)^33^. EI quantifies epileptogenicity by combining the high-/low-frequency energy ratio with the latency of the frequency transition relative to the first structure involved in the fast discharge. cEI extends this measure by incorporating directed functional connectivity (out-degree), enabling identification of leading epileptogenic nodes. SEEG contacts were classified as epileptogenic zone network (EZN; EI ≥ 0.4 and/or cEI ≥ 0.65), propagation zone network (PZN; 0.1 ≤ EI < 0.4 and/or 0.3 ≤ cEI < 0.65) or non-involved zone (NIZ; EI < 0.1 and/or cEI < 0.1)^34^. When multiple hippocampal-sampling contacts were present, classification was based on the maximum EI/cEI value across contacts and ictal events. As EI and cEI primarily capture seizure onset and early propagation, visual SEEG inspection complemented PZN classification in cases of late hippocampal involvement.

To quantify postictal signal dynamics, SEEG signal complexity was assessed using permutation entropy. PE measures the distribution of ordinal patterns derived from the relative ordering of values within a time series^35^. For each time window, patterns are defined according to an embedding dimension (D) and time delay (τ) and their probabilities (p(π)) are used to compute entropy: 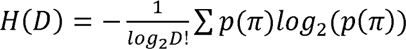 (details in Bratu et al.^22^). PE values range from 0 (deterministic, regular signals) to 1 (stochastic, random signals). SEEG PE analysis was performed using an in-house MATLAB plug-in (details in Bratu et al.^19^). Parameters were set to D = 3 samples and τ = 1 sample, using a 5-second sliding window with 2.5-second overlap, as in previous work.^19,22,23,25,36–38^ PE was computed for peri-ictal periods of spontaneous and stimulation-induced seizures. For quantification of the postictal period, as per our seminal work^19^, recovery of baseline signal complexity (“come-back point”) was defined as ≥ 25 consecutive seconds (10 overlapping windows) during which mean PE returned within ± 2 standard deviations of the preictal baseline. The interval between ictal offset and the “come-back point” defined the Postictal Alteration Time (postictal period)^19,23^. As PE operates on samples, all recordings were down-sampled to 256 Hz before PE quantification to account for heterogeneous sampling rates (256/512/1024/2048 Hz). A 30-minute baseline preceding spontaneous ictal activity or stimulation sessions was used. When ictal events occurred in close succession, the baseline preceding the first occurrence was retained, unless hippocampal signal complexity had fully recovered before the subsequent ictal event, allowing definition of a new baseline. Hippocampal ictal onset was defined as the earliest SEEG change within a sustained rhythmic discharge across bipolar hippocampal derivations and ictal offset as the cessation of ictal activity in the last involved derivation.^39^

### Seizure-related burden metrics

All ictal events between the 30-minute and one-week memory assessments were screened, including electro-clinical seizures and subclinical ictal discharges with clear non-interictal spatio-temporal evolution (e.g. changes in rhythmicity of interictal discharges). Only ictal events involving the hippocampus were retained for burden computation. For each event, the duration between hippocampal ictal onset and offset was computed and summed across ictal events to yield the cumulative hippocampal ictal burden (c-HipIC). In parallel, the maximum PAT observed across all hippocampal contacts **(Fig. 2)** was retained and summed across events to yield the cumulative hippocampal postictal burden (c-HipPIC). The cumulative hippocampal seizure-related burden (c-HipSZB) was defined as the sum of c-HipIC and c-HipPIC. All seizure-related burden metrics were computed separately for each hippocampus (left and right).

**Figure 2.**
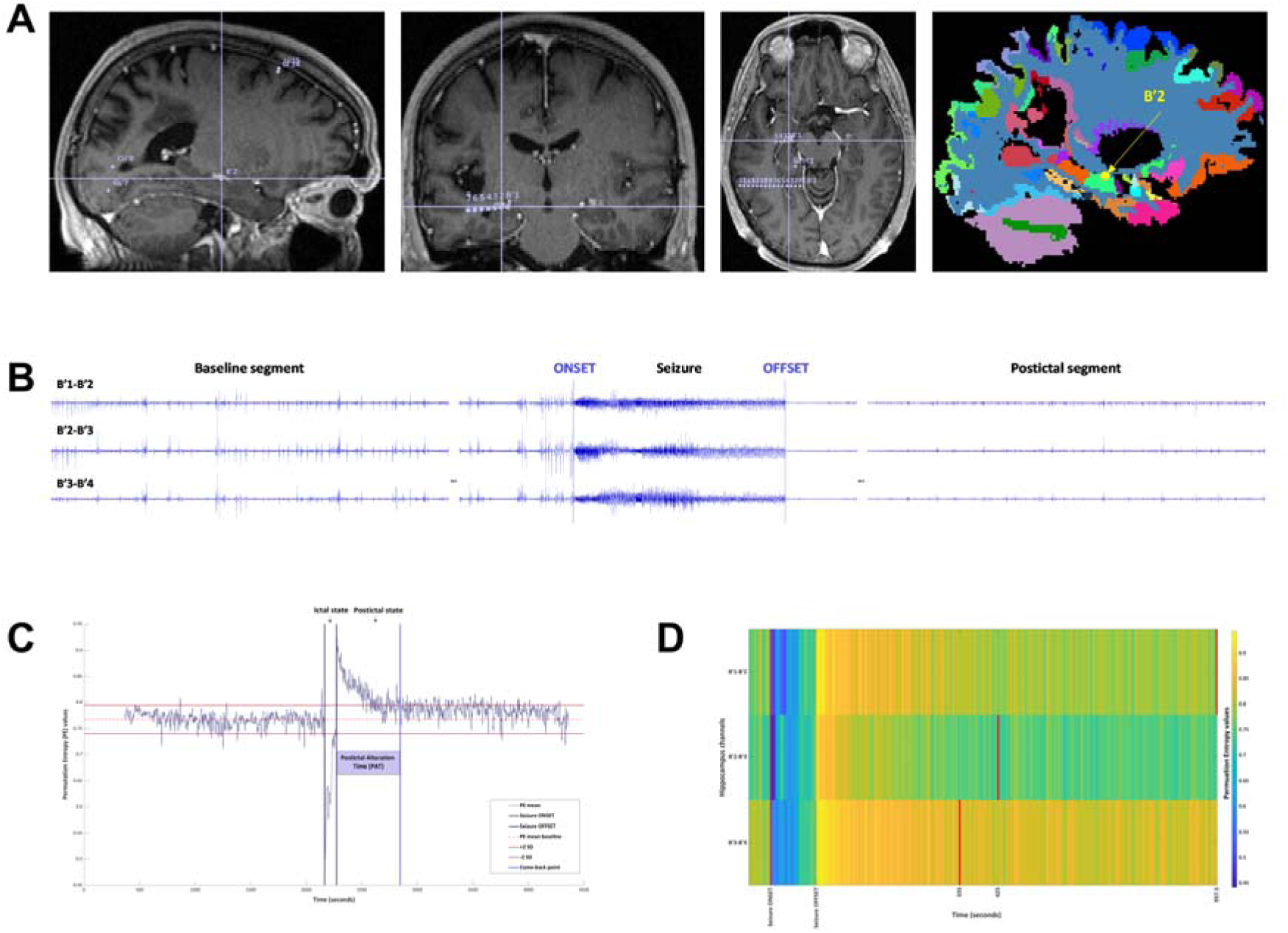
Hippocampal seizure-related burden metrics: anatomical-clinical and electrophysiological analysis pipeline. **(A)** Anatomical localization of hippocampal stereo-electroencephalography (SEEG) electrode contacts, reconstructed by co-registering the patient’s pre-SEEG implantation 3D T1-weighted cerebral MRI scan with the post-SEEG implantation cerebral CT scan. The first three panels display sagittal, coronal, and axial slices from the pre-SEEG implantation MRI, showing hippocampal sampling contacts and highlighting the B′2 contact (crosshairs). The fourth panel shows a sagittal slice from the pre-SEEG implantation cerebral MRI of the same patient, with left hemisphere parcellation based on the Virtual Epileptic Patient brain atlas projected into the patient’s native MRI space, further localizing B′2 within the hippocampus (highlighted by the yellow dot and arrow). **(B)** SEEG traces from left hippocampal bipolar derivations (B′1–B′2, B′2–B′3, B′3–B′4), illustrating baseline, ictal and postictal segments. Seizure onset and offset are marked by blue vertical bars. **(C)** Complexity dynamics of hippocampal electrical activity, captured by all hippocampal SEEG sampling contacts and quantified using permutation entropy (PE). The y-axis represents PE values and the x-axis corresponds to time in seconds. Vertical bars indicate seizure onset and termination (grey bars) and the “come-back point” (purple vertical bar)—defined as the moment at which the mean PE value stabilizes within two standard deviations of the preictal baseline for at least 25 consecutive seconds. The interval between seizure onset and termination defines the “ictal state”, while the interval from seizure termination to the “come-back point” defines the “postictal state”, also termed Postictal Alteration Time (PAT) in PE analysis. **(D)** Heatmap illustrating PE dynamics across the same hippocampal bipolar derivations as in (B). The colour gradient represents the PE spectrum, from dark blue (low entropy, high organization; minimum = 0) to bright yellow (high entropy, low organization; maximum = 1). The tall dark red vertical bar marks seizure onset, while short bright red vertical bars indicate the timing of the “come-back point” for each channel. The y-axis shows bipolar SEEG channels and the x-axis shows time in seconds.

### Hemispheric dominance

Language processing hemispheric organization was evaluated on a case-by-case basis using a multimodal framework integrating non-invasive and invasive assessments^23^. Initial lateralisation relied on non-invasive indicators, including ictal and postictal electro-clinical correlations from video-EEG recordings, relevant clinical features such as handedness and standardized age-appropriate neuropsychological evaluation. Language fMRI findings were incorporated within this non-invasive assessment when available. Lateralisation was further refined using data obtained during video-SEEG monitoring. This included analysis of electro-clinical correlations from video-SEEG recordings, assessment of auditory evoked potentials to phonemic contrasts (/ba/ vs /pa/)^40^ and functional mapping of language-eloquent regions through direct electrical stimulation.^41^ Task-related gamma-band activity during picture naming was also used to support identification of language-relevant cortical areas.^42^ When pre-operative findings remained inconclusive, post-operative language outcome was used to inform final classification, with preserved language function after epilepsy surgery supporting the inferred dominant hemisphere.

### Statistical analysis

Statistical analyses were conducted using GraphPad Prism (v10.6.1), MATLAB (v2023b) and Jamovi (v2.7.24) for Windows. Exploratory correlations assessed relationships between c-HipIC, c-HipPIC and c-HipSZB in the non-dominant-hemisphere hippocampus and visual memory performance and in the dominant-hemisphere hippocampus and verbal memory. As most variables violated normality (Shapiro–Wilk p < 0.05) Spearman’s rank correlation was used. These exploratory analyses were not corrected for multiple comparisons. As c-HipIC, c-HipPIC and c-HipSZB exhibited marked positive skewness and heteroscedasticity (range <100 to >60,000 seconds), a base-10 logarithmic transformation (Y = log (Y)) was applied to stabilise variance and limit the influence of extreme values. Memory performance at the 30-minute and one-week delays was expressed as percentages. One-week retention was defined as: 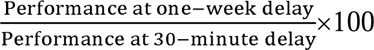. Associations identified in exploratory analyses were further examined using targeted univariate regression. Multivariate models were informed by univariate results and clinical relevance. Covariates **(Supplementary Table 1)** were tested in separate models: number of anti-seizure medications (ASMs), epilepsy duration, years of education and interictal spike rates (spikes/min; quantified during wakefulness and sleep (Lambert et al.^14^)). Exposure to ASMs with known prominent cognitive side effects (e.g., topiramate^43^, benzodiazepines, and barbiturates^44^), hippocampal EZN involvement and lesion status were modelled as binary variables (presence/absence). Histopathological findings were used when available; otherwise, lesion status was based on MRI. Given the limited sample size, models were restricted to few predictors to avoid overfitting. Multiple comparisons were controlled using false discovery rate (FDR, Benjamini–Hochberg). Statistical significance was set at α = 0.05.

## Results

### Patient characteristics

Patient characteristics are summarized in **Table 1**. Twenty patients (nine female, eleven male) were included. Mean age at seizure onset was 15.2 ± 11.4 years (range: 0.1–40) and mean age at SEEG was 32 ± 12 years (range: 14–56). EZN was bilateral in five patients and unilateral in fifteen (six right-sided, nine left-sided). Eighteen patients had a temporal lobe-involving EZN. Among the eleven patients with TLE, EZN was mesial in eight, lateral in two and mesial–lateral in one. Following SEEG evaluation, fourteen patients underwent curative-aim resective epilepsy surgery; five were contraindicated for surgery due to EZN extent and/or functional risk and one patient postponed surgery. Among operated patients, Engel class I outcome^45^ was achieved in 64% at last follow-up. Histopathological examination revealed non-specific findings in six cases (43%). Language dominance was left-sided in eighteen patients and right-sided in two. Patients received a mean of 2.8 ± 0.8 ASMs (range: 1–4), with six patients (30%) receiving medications associated with prominent cognitive side effects. Mean education was 12 ± 3.1 years (range: 5–17). No patient had active psychiatric comorbidity at SEEG **(Supplementary Table 1)**.

**Table 1.** Demographic, clinical and surgical characteristics of the study cohort.

| ID | Sex | EZN side | EZN localization/<br>temporal subtype | Resective<br>epilepsy surgery | Engel outcome<br>class at last<br>follow-up | Hemispheric<br>language<br>dominance |
| --- | --- | --- | --- | --- | --- | --- |
| P1 | F | R | T/mesial | Yes | I | L |
| P2 | M | B | T/mesial | Contraindicated | NA | L |
| P3 | F | B | Multilobar | Contraindicated | NA | L |
| P4 | F | R | T/mesial | Postponed | Non-I | L |
| P5 | M | R | T/mesial | Yes | I | L |
| P6 | M | L | F | Yes | I | L |
| P7 | M | R | F-T | Yes | I | L |
| P8 | M | L | T/mesial | Yes | I | R |
| P9 | M | B | T | Contraindicated | NA | L |
| P10 | M | L | F-T | Yes | Non-I | L |
| P11 | F | R | F-I | Yes | Non-I | L |
| P12 | M | L | T/mesial | Yes | I | L |
| P13 | F | R | T/lateral | Yes | NO | L |
| P14 | M | L | T/mesial | Yes | I | L |
| P15 | F | L | T/lateral | Yes | I | L |
| P16 | F | L | T/mesial | Yes | Non-I | L |
| P17 | F | B | T | Contraindicated | NA | L |
| P18 | F | L | T/mesial | Yes | I | L |
| P19 | M | L | T-P-O | Yes | Non-I | R |
| P20 | M | L | T/mesial-lateral | Contraindicated | NA | L |

F, female; M, male; SEEG, stereo-electroencephalography; EZN, epileptogenic zone network; R, right; L, left; B, bilateral; T, temporal; F, frontal; I, insular; P, parietal; O, occipital; M, mesial; ML, mesial-lateral; HS, hippocampal sclerosis; DNET, dysembryoplastic neuroepithelial tumour; FCD, focal cortical dysplasia; NA, not applicable; Non-I, Engel class II–IV outcome.

### Hippocampal characteristics

Among the thirty implanted and analysed hippocampi (fifteen left, fifteen right), six showed macroscopic structural lesions **(Table 2)**. Four of these lesions were confirmed histopathologically, while two belonged to patients who were not eligible for resective surgery. Of the ten hippocampi included in surgical resections, histopathology was positive in six cases: hippocampal sclerosis (3), gliosis (1), dysembryoplastic neuroepithelial tumour (1) and ganglioglioma (1). Fifteen implanted hippocampi belonged to the language-dominant hemisphere and fifteen to the non-dominant hemisphere. Nineteen hippocampi were classified as EZN (eight left, eleven right) and six as PZN (three left, three right). In the left hippocampus, spike rates averaged 5.1 ± 9.7 spikes/min during wakefulness (range: 0–28.1) and 15.1 ± 14.5 spikes/min during sleep (range: 1.2–44.2). In the right hippocampus, spike rates averaged 1.2 ± 1.4 spikes/min during wakefulness (range: 0–4.6) and 13.7 ± 10.7 spikes/min during sleep (range: 1.9–39.1).

**Table 2.** Hippocampal electro-clinical characteristics and memory testing.

| ID | SEEG sampling | MRI lesion/<br>Side | Resection/<br>pathology | Epileptogenicity<br>level | Ictal<br>events | Verbal testing | Visual testing |
| --- | --- | --- | --- | --- | --- | --- | --- |
| P1 | R | GGL | Yes/GGL | EZN | 2 | RL/RI, WMS* | DMS, WMS** |
| P2 | B | No | NA | EZN | 6 | RL/RI, WMS* | DMS, WMS** |
| P3 | B | No | NA | L – PZN<br>R - EZN | 4 | RL/RI, WMS* | DMS, WMS** |
| P4 | B | HS/R | NA | L – NIZ<br>R - EZN | 3 | RL/RI, WMS* | DMS, WMS** |
| P5 | R | No | Yes/non-specific | EZN | 4 | RL/RI | DMS, WMS** |
| P6 | L | No | No | NIZ | 1 | RL/RI, WMS* | DMS, WMS** |
| P7 | B | No | Yes/non-specific | L – NIZ<br>R - EZN | 2 | RL/RI, WMS** | DMS, WMS** |
| P8 | L | HS/L | Yes | EZN | 29 |  | DMS, WMS** |
| P9 | B | No | NA | L – PZN<br>R - EZN | 8 | RL/RI, WMS* | DMS, WMS** |
| P10 | B | No | Yes/non-specific | L – EZN<br>R - PZN | 3 | RL/RI, WMS* | DMS, WMS** |
| P11 | R | No | No | NIZ | 0 | RL/RI, WMS* | DMS, WMS** |
| P12 | B | DNET/L | Yes/DNET | EZN | 28 | WMS* | DMS, WMS** |
| P13 | B | No | No | L – NIZ<br>R - EZN | 7 | RL/RI, WMS* | DMS, WMS** |
| P14 | L | No | Yes/HS | EZN | 39 | RL/RI |  |
| P15 | L | No | No | EZN | 3 |  | DMS |
| P16 | R | No | Yes/HS | EZN | 40 | RL/RI | DMS |
| P17 | B | No | NA | EZN | 5 | RL/RI, WMS* | DMS, WMS** |
| P18 | B | Radionecrosis/L | Yes/Gliosis | L – EZN<br>R - PZN | 160 | RL/RI | DMS |
| P19 | R | No | Yes/non-specific | NIZ | 3 | RL/RI, WMS* | DMS, WMS** |
| P20 | L | Malformation and<br>malrotation/L | NA | PZN | 1 | RL/RI | DMS, WMS** |

Hip, hippocampus; R, right; L, left; B, bilateral; MRI, magnetic resonance imaging; GGL, Ganglioglioma; HS, hippocampal sclerosis; NA, not applicable; EZN, epileptogenic zone network; PZN, propagation zone network; NIZ, non-involved zone networks; RL/RI-16, Free and Cued Selective Reminding Test; WMS-III\*, Logical Memory subtest of the Wechsler Memory Scale—Third Edition; WMS-III\*\*, Visual Reproduction (Abstract Drawings) subtest of the WMS-III; DMS-48, Delayed Matching-to-Sample task.

### Seizure burden and memory assessment

Between the two memory testing sessions, 365 ictal events were identified. Fourteen were electrical stimulation-induced (eight patients), whereas 351 occurred spontaneously. The mean number of ictal events per patient was 18.3 ± 35.7 (range: 1–160) **(Table 2)**. Seventeen events were focal to bilateral tonic-clonic seizures (six patients).

Of the 365 events, 357 were assessable for hippocampal involvement, while eight were unavailable due to data storage issues. Among assessable events, 348 involved at least one hippocampus. 250 ictal events were retained for dominant-hemisphere hippocampal analyses (mean: 16.7 ± 39.5 per patient; range: 1–160) and 135 for non-dominant-hemisphere hippocampus analyses (mean: 9 ± 11 per patient; range: 0–40). Verbal memory assessments were available in eighteen patients (RL/RI-16: 17; WMS-III-LM: 13) and visual memory assessments were available in nineteen patients (DMS-48: 19; WMS-III-VR: 16).

### Hippocampal seizure-related burden metrics and memory scores at one-week delay – univariate analyses

Higher seizure-related burden metrics in the dominant hippocampus were consistently associated with poorer verbal memory performance and retention at the one-week delay. Exploratory Spearman correlation analyses revealed significant negative associations between c-HipIC, c-HipPIC and c-HipSZB and several verbal memory outcomes: RL and RT (RL/RI-16) and free and cued WMS-III-LM recall **(Supplementary Table 2)**. These findings were confirmed by univariate linear regression analyses **(Table 3)**, demonstrating robust negative associations (p < 0.05). Higher c-HipIC was associated with lower RT performance and retention (β = −26.66, R² = 0.44 and β = −27.46, R² = 0.48) **(Fig. 3)**, as well as with poorer WMS-III-LM free and cued recall performance (β = −45.33, R² = 0.70 and β = −13.29, R² = 0.52) and reduced free recall retention (β = −59.04, R² = 0.67). Similarly, higher c-HipPIC was associated with lower RL and RT performance (β = −12.49, R² = 0.34 and β = −19.83, R² = 0.48) and poorer WMS-III-LM free recall performance (β = −19.09, R² = 0.51). The strongest effects were observed for c-HipSZB. Higher c-HipSZB was associated with reduced RL and RT performance (β = −15.09, R² = 0.37 and β = −25.04, R² = 0.57), poorer WMS-III-LM free recall performance (β = −24.52, R² = 0.57) and reduced RT retention (β = −23.88, R² = 0.53).

**Table 3.** Univariate linear regressions between dominant-hemisphere hippocampal seizure-related burden metrics and verbal memory outcomes.

| Outcome | Burden type | Memory test | $\beta$ (slope) | 95% CI ($\beta$) | R<sup>2</sup> | FDR-p | N |
| --- | --- | --- | --- | --- | --- | --- | --- |
| **1-week performance** | c-HipIC | RL | -13.35 | -31.15 to 4.45 | 0.20 | 0.127 | 13 |
|  |  | RT | **-26.66** | **-46.50 to -6.82** | **0.44** | **0.022** | **13** |
|  |  | HIST R | **-45.33** | **-67.82 to -22.85** | **0.70** | **0.017** | **11** |
|  |  | HIST R C | **-13.29** | **-22.85 to -3.74** | **0.52** | **0.022** | **11** |
|  | c-HipPIC | RL | **-12.49** | **-24.07 to -0.92** | **0.34** | **0.049** | **13** |
|  |  | RT | **-19.83** | **-33.59 to -6.08** | **0.48** | **0.022** | **13** |
|  |  | HIST R | **-19.09** | **-33.08 to -5.10** | **0.51** | **0.022** | **11** |
|  |  | HIST R C | -4.74 | -10.52 to 1.04 | 0.28 | 0.106 | 11 |
|  | c-HipSZB | RL | **-15.09** | **-28.15 to -2.04** | **0.37** | **0.041** | **13** |
|  |  | RT | **-25.04** | **-39.46 to -10.63** | **0.57** | **0.017** | **13** |
|  |  | HIST R | **-24.52** | **-40.54 to -8.49** | **0.57** | **0.022** | **11** |
|  |  | HIST R C | -5.98 | -12.93 to 0.97 | 0.30 | 0.100 | 11 |
| **1-week retention** | c-HipIC | RL | -14.82 | -37.41 to 7.78 | 0.16 | 0.236 | 13 |
|  |  | RT | **-27.46** | **-46.36 to -8.55** | **0.48** | **0.034** | **13** |
|  |  | HIST R | **-59.04** | **-89.99 to -28.09** | **0.67** | **0.023** | **11** |
|  |  | HIST R C | -8.48 | -21.91 to 4.96 | 0.18 | 0.236 | 11 |
|  | c-HipPIC | RL | -13.39 | -28.64 to 1.85 | 0.25 | 0.159 | 13 |
|  |  | RT | -18.36 | -32.68 to -4.03 | 0.42 | 0.050 | 13 |
|  |  | HIST R | -16.88 | -40.24 to 6.47 | 0.23 | 0.205 | 11 |
|  |  | HIST R C | -0.96 | -8.22 to 6.30 | 0.01 | 0.771 | 11 |
|  | c-HipSZB | RL | -16.56 | -33.74 to 0.62 | 0.29 | 0.138 | 13 |
|  |  | RT | **-23.88** | **-38.75 to -9.02** | **0.53** | **0.028** | **13** |
|  |  | HIST R | -22.11 | -49.91 to 5.70 | 0.26 | 0.181 | 11 |
|  |  | HIST R C | -1.54 | -10.35 to 7.28 | 0.02 | 0.767 | 11 |

RL: Free recall score of the French Free and Cued Selective Reminding Test (RL/RI-16); RT: Total recall score (RL/RI-16; free recall + cued recall); HIST R: Free recall score on the Logical Memory subtest of Wechsler Memory Scale—Third Edition (WMS-III); HIST R C: Cued recall score on the Logical Memory subtest (WMS-III); c-HipIC: Cumulative hippocampal ictal burden (sum of ictal durations across seizures); c-HipPIC: Cumulative hippocampal postictal burden (sum of Postictal Alteration Time, postictal durations, across seizures); c-HipSZB: Cumulative hippocampal seizure-related burden (c-HipIC + c-HipPIC); CI: Confidence interval; R<sup>2</sup>: Coefficient of determination; N: Number of pairs.

**Figure 3.**
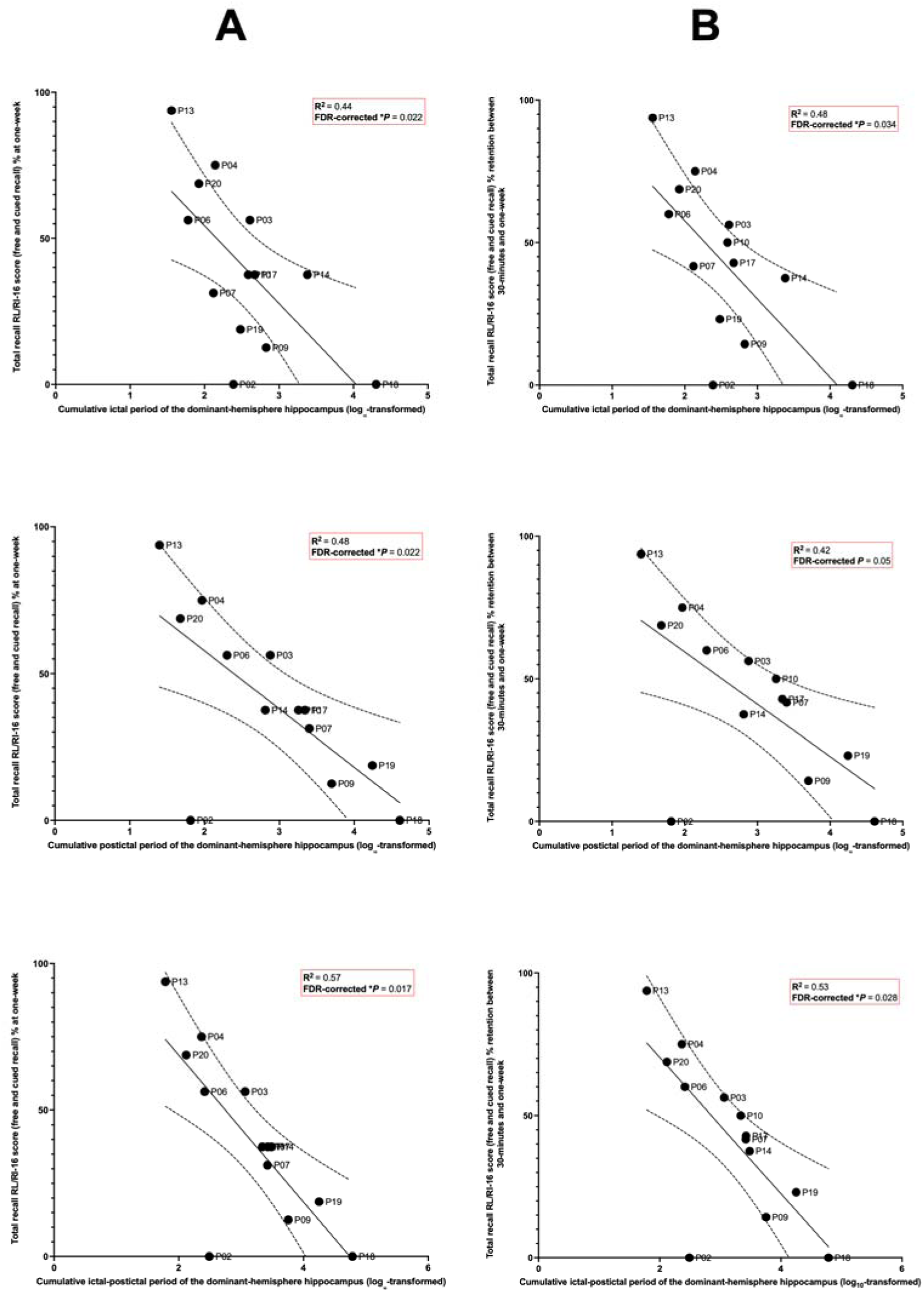
Associations between dominant-hemisphere hippocampal seizure-related burden metrics and verbal memory performance. Scatterplots illustrate univariate linear relationships between dominant-hemisphere cumulative hippocampal seizure-related burden metrics and total recall (free and cued) score on the French Free and Cued Selective Reminding Test (RL/RI-16). Column A shows associations with RT at the one-week delay, whereas column B shows associations with RT retention between the 30-minute and one-week assessments. Rows correspond to cumulative dominant-hemisphere hippocampal ictal burden, cumulative dominant-hemisphere hippocampal postictal burden and cumulative dominant-hemisphere hippocampal seizure-related burden (ictal-postictal). Burden metrics are expressed as logLL-transformed cumulative durations (original values in seconds). Each dot represents one patient; patient identifiers are indicated. Solid lines depict fitted linear regression slopes, with the two dotted lines indicating the 95% confidence interval around the regression estimate. Coefficients of determination (R²) and corresponding FDR-corrected p values are reported within each panel.

In contrast, seizure-related burden metrics in the non-dominant hippocampus showed no robust associations with visual memory, either for one-week performance or retention. Exploratory analyses revealed isolated associations with WMS-III-VR (positive for c-HipPIC and c-HipSZB for performance; negative for c-HipPIC for retention), but these were not reproduced across other visual memory outcomes nor observed consistently across burden metrics **(Supplementary Table 3)**. Several associations showed weak or opposite-direction trends. Given the lack of consistency, these associations were not analysed further.

### Hippocampal seizure-related burden metrics and memory scores at one-week delay – multivariate analyses

Among verbal memory outcomes, RT was prioritised for multivariate analyses as it showed the most robust association with hippocampal seizure-related burden metrics. Multivariate linear regression models were therefore constructed to examine the extent to which dominant-hemisphere hippocampal burden metrics explain variance in RT performance and retention at one week, adjusting separately for clinical and electrophysiological covariates **(Tables 4–5)**. Higher burden metrics were consistently associated with poorer RT performance and retention at one week, explaining a substantial proportion of RT variance, with associations generally remaining significant after covariate adjustment. Models including c-HipIC showed moderate explanatory power (adjusted R² = 0.33–0.45 for RT performance and 0.38–0.46 for RT retention), but associations were not consistently significant across covariate adjustments, particularly when accounting for hippocampal EZN involvement or spike rate during sleep. In contrast, models including c-HipPIC explained a greater proportion of variance in RT performance (adjusted R² = 0.38–0.52) and showed moderate performance in explaining RT retention (adjusted R² = 0.30–0.46), with associations remaining significant when adjusting for most covariates, except for spike rates (during sleep for performance and during either sleep or wakefulness for retention). Models including c-HipSZB consistently explained the largest proportion of variance (adjusted R² = 0.49–0.59 for RT performance and 0.44–0.53 for RT retention), remaining significant across all tested covariates for performance and losing significance only when adjusting for spike rate during sleep for retention. Standardized effect sizes were consistently large, supporting a robust relationship between seizure-related burden and both memory performance and retention. For RT performance, the best-performing models (p < 0.05) were obtained when adjusting for lesion status for c-HipIC (F(2,10) = 5.86; adjusted R² = 0.45), EZN involvement for c-HipPIC (F(2,10) = 7.66; adjusted R² = 0.53) and epilepsy duration for c-HipSZB (F(2,10) = 9.46; adjusted R² = 0.59). For retention, best-performing models were observed when adjusting for epilepsy duration for c-HipIC (F(2,10) = 6.10; adjusted R² = 0.46) and c-HipSZB (F(2,10) = 7.78; adjusted R² = 0.53) and number of ASMs for c-HipPIC (F(2,10) = 6.10; adjusted R² = 0.46).

**Table 4.** **Multivariate linear regression models examining the association between dominant-hemisphere hippocampal ictal (c-HipIC), postictal (c-HipPIC) and seizure-related (c-HipSZB) burden and total recall score (RT) at 1-week*\****

| Predictor | Covariate | $\beta$ (SE) | 95% CI | Standardized $\beta$ | p-value | Model F (df=2,10) | Adjusted R<sup>2</sup> | AIC |
| --- | --- | --- | --- | --- | --- | --- | --- | --- |
| **c-HipIC** | Number of ASMs | -26.64 (9.43) | [-47.70, -5.62] | -0.67 | 0.018 | 4.01 | 0.33 | 124 |
|  | Cognitive-ASM | -25.20 (9.65) | [-46.70, -3.70] | -0.63 | 0.026 | 4.27 | 0.35 | 123 |
|  | EZN involvement | -22.76 (11.90) | [-49.40, 3.84] | -0.57 | 0.086 | 4.22 | 0.35 | 123 |
|  | Epilepsy duration | -26.90 (8.81) | [-46.53, -7.27] | -0.67 | 0.012 | 5.34 | 0.42 | 122 |
|  | Spike rate (wakefulness) | -42.52 (18.23) | [-83.14, -1.90] | -1.06 | 0.042 | 4.87 | 0.39 | 122 |
|  | Spike rate (sleep) | -20.05 (13.49) | [-50.12, 10.01] | -0.50 | 0.168 | 4.38 | 0.36 | 123 |
|  | Years of education | -26.68 (9.48) | [-47.81, -5.55] | -0.67 | 0.018 | 3.98 | 0.33 | 124 |
|  | **Lesion** | **-34.70 (10.20)** | **[-57.40, -11.90]** | **-0.87** | **0.007** | **5.86** | **0.45** | **121** |
| **c-HipPIC** | Number of ASMs | -24.20 (6.21) | [-38.00, -10.30] | -0.84 | 0.003 | 7.59 | 0.52 | 119 |
|  | Cognitive-ASM | -20.30 (7.46) | [-36.90, -3.69] | -0.71 | 0.021 | 4.59 | 0.38 | 123 |
|  | **EZN involvement** | **-17.30 (5.87)** | **[-30.40, -4.26]** | **-0.60** | **0.014** | **7.66** | **0.53** | **119** |
|  | Epilepsy duration | -20.54 (5.93) | [-33.76, -7.33] | -0.72 | 0.006 | 6.77 | 0.49 | 120 |
|  | Spike rate (wakefulness) | -18.41 (8.15) | [-36.56, -0.25] | -0.64 | 0.047 | 4.66 | 0.38 | 123 |
|  | Spike rate (sleep) | -15.28 (8.39) | [-33.96, 3.40] | -0.53 | 0.098 | 5.24 | 0.41 | 122 |
|  | Years of education | -20.39 (6.49) | [-34.85, -5.93] | -0.71 | 0.010 | 4.96 | 0.40 | 122 |
|  | Lesion | -19.74 (6.57) | [-34.40, -5.10] | -0.69 | 0.013 | 4.60 | 0.38 | 123 |
| **c-HipSZB** | Number of ASMs | -27.40 (6.52) | [-41.96, -12.90] | -0.83 | 0.002 | 8.88 | 0.57 | 118 |
|  | Cognitive-ASM | -25.71 (7.68) | [-42.80, -8.60] | -0.78 | 0.007 | 6.69 | 0.49 | 120 |
|  | EZN involvement | -21.80 (6.74) | [-36.80, -6.76] | -0.66 | 0.009 | 8.82 | 0.57 | 118 |
|  | **Epilepsy duration** | **-25.45 (6.17)** | **[-39.21, -11.70]** | **-0.77** | **0.002** | **9.46** | **0.59** | **117** |
|  | Spike rate (wakefulness) | -26.98 (9.32) | [-47.75, -6.21] | -0.81 | 0.016 | 6.75 | 0.49 | 120 |
|  | Spike rate (sleep) | -22.10 (9.46) | [-43.17, -1.02] | -0.67 | 0.042 | 6.88 | 0.50 | 120 |
|  | Years of education | -25.27 (6.81) | [-40.43, -10.10] | -0.76 | 0.004 | 6.91 | 0.50 | 120 |
|  | Lesion | -25.54 (7.00) | [-41.10, -9.95] | -0.77 | 0.004 | 6.77 | 0.49 | 120 |

\*French Free and Cued Selective Reminding Test (RL/RI-16); RT: Total recall score (RL/RI-16; free recall + cued recall). Cognitive-ASM, ASM with known prominent cognitive side effects.

**Table 5.** Multivariate linear regression models examining the association between dominant-hemisphere hippocampal ictal (c-HipIC), postictal (c-HipPIC) and seizure-related (c-HipSZB) burden and verbal memory retention (RT) between 30-minute and 1-week delay*.

| Predictor | Covariate | $\beta$ (SE) | 95% CI | Standardized $\beta$ | p-value | Model F (df=2,10) | Adjusted R<sup>2</sup> | AIC |
| --- | --- | --- | --- | --- | --- | --- | --- | --- |
| **c-HipIC** | Number of ASMs | -27.43 (8.96) | [-47.4, -7.46] | -0.69 | 0.012 | 4.74 | 0.38 | 122 |
|  | Cognitive-ASM | -26.00 (9.18) | [-46.5, -5.55] | -0.66 | 0.018 | 4.99 | 0.40 | 122 |
|  | EZN involvement | -24.82 (11.50) | [-50.3, 0.70] | -0.63 | 0.055 | 4.78 | 0.39 | 122 |
|  | **Epilepsy duration** | **-27.69 (8.40)** | **[-46.40, -8.97]** | **-0.70** | **0.008** | **6.10** | **0.46** | **121** |
|  | Spike rate (wakefulness) | -43.08 (17.31) | [-81.65, -4.51] | -1.09 | 0.032 | 5.69 | 0.44 | 121 |
|  | Spike rate (sleep) | -22.61 (12.97) | [-51.51, 6.30] | -0.57 | 0.112 | 4.90 | 0.39 | 122 |
|  | Years of education | -27.44 (9.04) | [-47.57, -7.31] | -0.69 | 0.013 | 4.65 | 0.38 | 122 |
|  | Lesion | -34.00 (10.00) | [-56.3, -11.7] | -0.86 | 0.007 | 6.06 | 0.46 | 121 |
| **c-HipPIC** | **Number of ASMs** | **-22.70 (6.54)** | **[-37.27, -8.13]** | **-0.80** | **0.006** | **6.10** | **0.46** | **121** |
|  | Cognitive-ASM | -18.30 (7.77) | [-35.60, -0.98] | -0.65 | 0.040 | 3.62 | 0.30 | 124 |
|  | EZN involvement | -15.90 (6.22) | [-29.80, -2.04] | -0.56 | 0.029 | 6.00 | 0.46 | 121 |
|  | Epilepsy duration | -19.02 (6.31) | [-33.07, -4.97] | -0.67 | 0.013 | 5.16 | 0.41 | 122 |
|  | Spike rate (wakefulness) | -15.60 (8.39) | [-34.30, 3.11] | -0.55 | 0.093 | 3.88 | 0.32 | 123 |
|  | Spike rate (sleep) | -13.15 (8.67) | [-32.47, 6.17] | -0.46 | 0.160 | 4.34 | 0.36 | 123 |
|  | Years of education | -18.93 (6.76) | [-33.99, -3.88] | -0.67 | 0.019 | 3.96 | 0.33 | 123 |
|  | Lesion | -18.10 (6.79) | [-33.20, -2.96] | -0.64 | 0.024 | 3.77 | 0.32 | 124 |
| **c-HipSZB** | Number of ASMs | -26.40 (6.70) | [-41.32, -11.50] | -0.81 | 0.003 | 7.84 | 0.53 | 119 |
|  | Cognitive-ASM | -24.17 (7.93) | [-41.80, -6.50] | -0.74 | 0.012 | 5.69 | 0.44 | 121 |
|  | EZN involvement | -20.70 (7.02) | [-36.30, -5.06] | -0.63 | 0.015 | 7.44 | 0.52 | 119 |
|  | **Epilepsy duration** | **-24.27 (6.48)** | **[-38.72, -9.83]** | **-0.74** | **0.004** | **7.78** | **0.53** | **119** |
|  | Spike rate (wakefulness) | -24.03 (9.65) | [-45.54, -2.52] | -0.73 | 0.032 | 5.69 | 0.44 | 121 |
|  | Spike rate (sleep) | -20.47 (9.73) | [-42.15, 1.20] | -0.63 | 0.062 | 5.96 | 0.45 | 121 |
|  | Years of education | -24.13 (7.01) | [-39.74, -8.52] | -0.74 | 0.006 | 5.97 | 0.45 | 121 |
|  | Lesion | -23.89 (7.26) | [-40.10, -7.72] | -0.73 | 0.008 | 5.69 | 0.44 | 121 |

\*French Free and Cued Selective Reminding Test (RL/RI-16); RT: Total recall score (RL/RI-16; free recall + cued recall). Cognitive-ASM, ASM with known prominent cognitive side effects.

## Discussion

This study introduces the concept of cumulative hippocampal seizure-related burden as a quantitative marker linking epileptic activity to long-term memory consolidation. We show that this burden explains a substantial proportion of variance in long-term verbal memory outcomes over a one-week delay and is the strongest and most consistent multivariate predictor of both performance and retention. Among burden metrics, cumulative ictal–postictal burden (c-HipSZB) showed the most robust associations in covariate-adjusted models, whereas ictal and postictal burdens considered separately were less consistent, particularly when accounting for interictal spike rates. These findings indicate that the cumulative temporal footprint of seizure-related hippocampal dysfunction is a key determinant of long-term memory outcome in focal drug-resistant epilepsy.

Unlike traditional metrics (e.g., seizure count or frequency), c-HipSZB captures the cumulative duration of hippocampal dysfunction by integrating cumulative ictal and postictal periods across all hippocampus-involving ictal events, including electroclinical seizures and subclinical ictal discharges with clear spatiotemporal evolution. Such cumulative approaches are essential for understanding the pathophysiology of progressive cognitive comorbidities in epilepsy, especially memory^46^.

A key finding is that hippocampal seizure-related burden was robustly associated with recall, but not with recognition. This dissociation aligns with the hippocampus-centric focus of the present work and established models of episodic memory, in which the hippocampus primarily supports recall, whereas recognition depends predominantly on perirhinal and entorhinal regions^47^. Accordingly, selective hippocampal dysfunction is expected to disproportionately impair recall while relatively sparing recognition^48^. This is supported by empirical evidence: patients with focal perirhinal or anteromedial temporal lesions (sparing the hippocampus) show impaired recognition with relatively preserved recall, whereas patients with isolated hippocampal damage show severe recall deficits with relative preservation of recognition^49^. Consistent with this hippocampal recall-specific vulnerability, we also observed differential sensitivity across recall paradigms: burden metrics were robustly associated with list-based verbal recall (RL/RI-16), whereas associations with story recall were less consistent. Whether list-learning or narrative memory tasks are more sensitive to hippocampal dysfunction remains debated.^50^

Our findings provide only partial support for material-specific lateralization of hippocampal function. Cumulative seizure-related burden in the dominant-hemisphere hippocampus showed robust associations with verbal memory outcomes, consistent with literature linking the left hippocampus with verbal memory.^5,6,51,52^ In contrast, no robust associations were observed between non-dominant-hemisphere hippocampal burden and visual memory. This asymmetry should be interpreted cautiously. First, hippocampal material specificity does not invariably map onto language dominance in terms of hemispheric dominance.^53–56^ Second, visual memory assessment relied predominantly on recognition-based tasks, whereas hippocampal burden preferentially affected recall. Third, visual memory likely depends on more bilateral and distributed medial temporal networks than verbal memory. Accordingly, a unilateral hippocampal burden approach may not fully capture the network-level mechanisms underlying visual memory performance.^57^

Prior studies have explored various predictors of memory impairment in epilepsy, including age at onset, seizure frequency, epilepsy duration, psychiatric comorbidities, MRI-visible lesions and hippocampal volumetry.^3,13,58,59^ In contrast, intracranial EEG studies—allowing direct quantification of epileptic activity within memory-relevant structures—remain comparatively scarce, despite their potential to provide more direct evidence than traditional clinical variables. Aykan et al.^60^ showed in SEEG that working memory performance negatively correlates with frontotemporal theta-band connectivity, itself linked to mesial temporal spike rate and theta power. Moreover, Lambert et al.^14^ demonstrated that long-term verbal memory consolidation over a one-week delay is impaired in TLE, with seizure number and hippocampal interictal activity during NREM sleep emerging as detrimental factors. In our models, cumulative hippocampal seizure-related burden metrics remained significantly associated with long-term verbal memory and retention after adjustment for multiple covariates (ASMs, epilepsy duration, education, hippocampal lesion status, EZN involvement and interictal spike rate). Overall model fits were robust; however, inclusion of certain covariates, especially spike rates, attenuated the strength or statistical significance of these associations in some models. Importantly, this does not imply that clinical variables are irrelevant to cognitive outcome, but rather that their influence may be mediated through cumulative hippocampal seizure-related dysfunction.

This monocentric study has several limitations. The relatively small sample size limits generalisability, constrains multivariate modelling, and reduces sensitivity to detect subtle effects. Although hemispheric dominance was assessed using a comprehensive multimodal approach, misclassification cannot be excluded. In patients with long-standing epilepsy or structural brain lesions, language-memory networks may be reorganised over time.^56^ Furthermore, ASM dose modifications during SEEG monitoring may influence memory performance, but were not accounted for. Hippocampal lesion status was treated as a binary variable based on clinical MRI or histopathology, without accounting for lesion type, severity or volumetry, potentially limiting modelling of structural heterogeneity. Variability in clinical condition during SEEG also limited completion of the full memory assessment battery in all participants. Additional limitations are inherent to the SEEG evaluation, including hypothesis-driven implantation based on non-invasive data, limited spatial sampling and inter-individual variability in the brain regions explored. Finally, although alternative PE-derived metrics (e.g. maximal postictal deviation or integrative measures combining magnitude and duration) may capture complementary postictal dynamics, they rely on the early postictal phase—previously shown to correspond to maximal entropy disorganisation^22^—which was frequently contaminated by SEEG artefacts, limiting reliable and standardised extraction.

Despite these limitations, this study addresses the difficulty of quantifying cumulative seizure effects using reliable and physiologically grounded measures.^15^ By exploiting continuous SEEG recordings, we move beyond seizure counts to capture the cumulative duration of hippocampal dysfunction across ictal and postictal states. The association between hippocampal burden metrics and long-term verbal memory performance provides functional evidence that repeated seizure-related disruptions, considered as a temporal ictal-postictal continuum, impact cognitive outcomes in focal epilepsy. Importantly, this work extends our prior efforts to conceptualise the postictal state as a quantifiable contributor to epilepsy-related morbidity.^23,61,62^

These findings support the relevance of long-term electrophysiological monitoring approaches to quantify seizure-related burden. Implantable neuromodulation and sensing devices with chronic recording capabilities offer new opportunities to track hippocampal dysfunction over extended timescales. Coupling such recordings with longitudinal memory assessment protocols targeting consolidation processes may enable more sensitive monitoring of cognitive trajectories and treatment effects in focal epilepsy.

## Data availability

The AnyWave software and dedicated plug-ins (Epileptogenicity Index – EI and Connectivity Epileptogenicity Index - cEI) are freely available at https://meg.univ-amu.fr/AnyWave/download.html and https://meg.univ-amu.fr/doku.php?id=plugins:ei. The Virtual Epileptic Patient (VEP) brain atlas parcellation is available open source at https://ins-amu.fr/vep-atlas. The Permutation Entropy plug-in, the Postictal Alteration Time code and the datasets generated and analysed during the current study are available from the corresponding author upon reasonable request.

## Author contributions

**I.F.B.**: Conceptualization; data curation; formal analysis; investigation; methodology; project administration; software; validation; visualization; writing—original draft; writing—review and editing. **I.L.**: Conceptualization; data curation; investigation; methodology; writing—review and editing. **O.F.**: Conceptualization; methodology; investigation; writing—review and editing. **S.M.V.**: Software; visualization; writing—review and editing. **A.T.**: Conceptualization; methodology; investigation; project administration; supervision; validation; writing—review and editing. **F.B.:** Conceptualization; data curation; funding acquisition; investigation; methodology; project administration; resources; supervision; validation; writing—review and editing.

## Acknowledgements

We thank Bernard Giusiano and Capucine Rodet for their valuable insight regarding the statistical analyses. We thank Prof Didier Scavarda and Dr Romain Carron and their teams for the surgical management of included patients; Dr Julia Makhalova, Dr Lagarde Stanislas, Dr Francesca Pizzo, Dr Francesca Bonini, Dr Aileen McGonigal, Dr Lisa Vaugier, Dr Sandrine Aubert, Dr Nathalie Villeneuve, Dr Anne Lepine and their teams for the clinical management of some included patients. The project leading to this publication has received support from the French government under the “France 2030” investment plan managed by the French National Research Agency (reference: ANR-16-CONV000X/ANR-17-EURE-0029), from Excellence Initiative of Aix-Marseille University - A*MIDEX (AMX-19-IET-004) and from the VESTISELF project (Reference: ANR-19-CE37-0027).

