## Supplementary Table 3 for "Cumulative hippocampal seizure-related burden impairs long-term memory consolidation in focal epilepsy"

| **Table 3. Exploratory Spearman correlations between non-dominant-hemisphere hippocampal seizure-related burden metrics and visual memory outcomes** | | | | | | |
| --- | --- | --- | --- | --- | --- | --- |
| **Outcome** | **Burden type** | **Memory test** | **Spearman ρ** | **95% CI** | **p-value*** | **N** |
| **1-week performance** | c-HipIC | DMS rec | −0.35 | −0.75 to 0.23 | 0.212 | 14 |
|  |  | DMS pos | −0.11 | −0.61 to 0.46 | 0.716 | 14 |
|  |  | DMS dist | 0.03 | −0.52 to 0.56 | 0.927 | 14 |
|  |  | Rappel des | −0.21 | −0.71 to 0.43 | 0.510 | 12 |
|  |  | Rec vis cib | 0.30 | −0.35 to 0.75 | 0.339 | 12 |
|  |  | Rec vis dist | −0.47 | −0.83 to 0.16 | 0.128 | 12 |
|  | c-HipPIC | DMS rec | 0.05 | −0.50 to 0.58 | 0.857 | 14 |
|  |  | DMS pos | −0.20 | −0.67 to 0.38 | 0.483 | 14 |
|  |  | DMS dist | −0.03 | −0.56 to 0.52 | 0.921 | 14 |
|  |  | Rappel des | −0.27 | −0.74 to 0.38 | 0.400 | 12 |
|  |  | Rec vis cib | 0.65 | 0.10 to 0.90 | 0.026 | 12 |
|  |  | **Rec vis dist** | **−0.61** | **−0.88 to −0.03** | **0.041** | **12** |
|  | c-HipSZB | DMS rec | −0.14 | −0.64 to 0.43 | 0.624 | 14 |
|  |  | DMS pos | −0.18 | −0.66 to 0.40 | 0.532 | 14 |
|  |  | DMS dist | 0.05 | −0.50 to 0.58 | 0.858 | 14 |
|  |  | Rappel des | −0.23 | −0.72 to 0.42 | 0.481 | 12 |
|  |  | Rec vis cib | 0.58 | −0.01 to 0.87 | 0.051 | 12 |
|  |  | Rec vis dist | −0.56 | −0.86 to 0.05 | 0.065 | 12 |
| **1-week retention** | c-HipIC | DMS rec | −0.28 | −0.71 to 0.31 | 0.336 | 14 |
|  |  | DMS pos | −0.23 | −0.69 to 0.36 | 0.436 | 14 |
|  |  | DMS dist | −0.08 | −0.60 to 0.49 | 0.789 | 14 |
|  |  | Rappel des | −0.10 | −0.67 to 0.55 | 0.773 | 11 |
|  |  | Rec vis cib | 0.21 | −0.46 to 0.73 | 0.538 | 11 |
|  |  | Rec vis dist | −0.60 | −0.89 to 0.02 | 0.055 | 11 |
|  | c-HipPIC | DMS rec | 0.12 | −0.46 to 0.62 | 0.694 | 14 |
|  |  | DMS pos | −0.27 | −0.71 to 0.32 | 0.357 | 14 |
|  |  | DMS dist | −0.05 | −0.58 to 0.51 | 0.873 | 14 |
|  |  | Rappel des | −0.27 | −0.76 to 0.41 | 0.429 | 11 |
|  |  | Rec vis cib | 0.58 | −0.06 to 0.88 | 0.066 | 11 |
|  |  | **Rec vis dist** | **−0.85** | **−0.96 to −0.50** | **0.0014** | **11** |
|  | c-HipSZB | DMS rec | −0.05 | −0.58 to 0.51 | 0.870 | 14 |
|  |  | DMS pos | −0.31 | −0.73 to 0.28 | 0.281 | 14 |
|  |  | DMS dist | −0.02 | −0.56 to 0.53 | 0.935 | 14 |
|  |  | Rappel des | −0.17 | −0.71 to 0.50 | 0.630 | 11 |
|  |  | Rec vis cib | 0.53 | −0.12 to 0.86 | 0.095 | 11 |
|  |  | **Rec vis dist** | **−0.77** | **−0.94 to −0.31** | **0.0069** | **11** |

DMS rec: Visual recognition of targets on the Delayed Matching-to-Sample task (DMS-48); DMS pos: Spatial position accuracy for correctly recognised targets (DMS-48); DMS dist: Recognition of distractors (false alarms) on the DMS-48; Rappel des: Free recall score on the Visual Reproduction (Abstract Drawings) subtest of the Wechsler Memory Scale—Third Edition (WMS-III); Rec vis cib: Visual recognition of targets on the WMS-III Visual Reproduction recognition task; Rec vis dist: Recognition of distractors on the WMS-III Visual Reproduction recognition task; c-HipIC: Cumulative hippocampal ictal burden (sum of ictal durations across seizures); c-HipPIC: Cumulative hippocampal postictal burden (sum of Postictal Alteration Time, postictal durations, across seizures); c-HipSZB: Cumulative hippocampal seizure-related burden (c-HipIC + c-HipPIC); CI: Confidence interval; N: Number of pairs.

*p-values are uncorrected for multiple comparisons.
