## Supplementary Table 2 for "Cumulative hippocampal seizure-related burden impairs long-term memory consolidation in focal epilepsy"

| **Table 2. Exploratory Spearman correlations between dominant-hemisphere hippocampal seizure-related burden metrics and verbal memory outcomes** | | | | | | |
| --- | --- | --- | --- | --- | --- | --- |
| **Outcome** | **Burden type** | **Memory test** | **Spearman ρ** | **95% CI** | **p-value*** | **N** |
| **1-week performance** | c-HipIC | RL | −0.43 | −0.80 to 0.18 | 0.147 | 13 |
|  |  | RT | −0.59 | −0.87 to −0.04 | 0.036 | 13 |
|  |  | RL RI REC | −0.33 | −0.75 to 0.29 | 0.267 | 13 |
|  |  | RL RI DIST | −0.48 | −0.82 to 0.12 | 0.101 | 13 |
|  |  | HIST R | −0.83 | −0.96 to −0.44 | 0.002 | 11 |
|  |  | HIST R C | −0.81 | −0.95 to −0.38 | 0.004 | 11 |
|  | c-HipPIC | RL | −0.58 | −0.86 to −0.03 | 0.040 | 13 |
|  |  | RT | −0.69 | −0.90 to −0.20 | 0.012 | 13 |
|  |  | RL RI REC | −0.42 | −0.80 to 0.19 | 0.151 | 13 |
|  |  | RL RI DIST | −0.31 | −0.74 to 0.31 | 0.300 | 13 |
|  |  | HIST R | −0.72 | −0.93 to −0.19 | 0.015 | 11 |
|  |  | HIST R C | −0.46 | −0.84 to 0.21 | 0.155 | 11 |
|  | c-HipSZB | RL | −0.71 | −0.91 to −0.24 | 0.009 | 13 |
|  |  | RT | −0.78 | −0.93 to −0.39 | 0.002 | 13 |
|  |  | RL RI REC | −0.30 | −0.74 to 0.32 | 0.312 | 13 |
|  |  | RL RI DIST | −0.34 | −0.76 to 0.28 | 0.259 | 13 |
|  |  | HIST R | −0.82 | −0.95 to −0.42 | 0.003 | 11 |
|  |  | HIST R C | −0.56 | −0.87 to 0.09 | 0.081 | 11 |
| **1-week retention** | c-HipIC | RL | −0.39 | −0.78 to 0.23 | 0.193 | 13 |
|  |  | RT | −0.67 | −0.90 to −0.18 | 0.014 | 13 |
|  |  | RL RI REC | −0.13 | −0.64 to 0.47 | 0.677 | 13 |
|  |  | RL RI DIST | −0.48 | −0.82 to 0.12 | 0.101 | 13 |
|  |  | HIST R | −0.95 | −0.99 to −0.81 | <0.0001 | 11 |
|  |  | HIST R C | −0.52 | −0.86 to 0.13 | 0.101 | 11 |
|  | c-HipPIC | RL | −0.53 | −0.84 to 0.05 | 0.066 | 13 |
|  |  | RT | −0.65 | −0.89 to −0.14 | 0.018 | 13 |
|  |  | RL RI REC | −0.10 | −0.63 to 0.49 | 0.740 | 13 |
|  |  | RL RI DIST | −0.31 | −0.74 to 0.31 | 0.300 | 13 |
|  |  | HIST R | −0.46 | −0.84 to 0.21 | 0.156 | 11 |
|  |  | HIST R C | −0.12 | −0.68 to 0.53 | 0.728 | 11 |
|  | c-HipSZB | RL | −0.66 | −0.89 to −0.15 | 0.017 | 13 |
|  |  | RT | −0.81 | −0.94 to −0.45 | 0.001 | 13 |
|  |  | RL RI REC | −0.00 | −0.57 to 0.56 | 0.996 | 13 |
|  |  | RL RI DIST | −0.34 | −0.76 to 0.28 | 0.259 | 13 |
|  |  | HIST R | −0.58 | −0.88 to 0.05 | 0.067 | 11 |
|  |  | HIST R C | −0.19 | −0.72 to 0.48 | 0.581 | 11 |

RL: Free recall score of the French Free and Cued Selective Reminding Test (RL/RI-16); RT: Total recall score (RL/RI-16; free recall + cued recall); RL RI REC: Recognition of targets on the RL/RI-16 recognition task; RL RI DIST: Recognition of distractors on the RL/RI-16 recognition task (false alarms); HIST R: Free recall score on the Logical Memory subtest of Wechsler Memory Scale—Third Edition (WMS-III); HIST R C: Cued recall score on the Logical Memory subtest (WMS-III); c-HipIC: Cumulative hippocampal ictal burden (sum of ictal durations across seizures); c-HipPIC: Cumulative hippocampal postictal burden (sum of Postictal Alteration Time, postictal durations, across seizures); c-HipSZB: Cumulative hippocampal seizure-related burden (c-HipIC + c-HipPIC); CI: Confidence interval; N: Number of pairs.

*p-values are uncorrected for multiple comparisons.
