## Supplementary Table 1 for "Cumulative hippocampal seizure-related burden impairs long-term memory consolidation in focal epilepsy"

| **Supplementary Table 1. Clinical and electrophysiological covariates** | | | | | | | | | |
| --- | --- | --- | --- | --- | --- | --- | --- | --- | --- |
| **ID** | **ASM Regimen** | **Number of ASMs** | **ASM with prominent cognitive**  **side effects** | **Psychiatric comorbidity** | **Years of education** | **Spike rate (wakefulness,**  **left hippocampus)** | **Spike rate**  **(sleep,**  **left hippocampus)** | **Spike rate**  **(wakefulness,**  **right hippocampus)** | **Spike rate**  **(sleep,**  **right hippocampus)** |
| P1 | LCS, LTG | 2 | No | No | 15 | – | – | 2.59 | 39.13 |
| P2 | CBZ | 1 | No | No | 11 | 0.13 | 9.23 | 0.03 | 11.33 |
| P3 | PER, LCS, LTG | 3 | No | No | 16 | 0.89 | 3.78 | 0.95 | 8.13 |
| P4 | LCS, CBZ | 2 | No | No | 9 | 0.00 | 3.57 | 0.77 | 26.68 |
| P5 | LCS, LEV | 2 | No | No | 12 | – | – | 0.67 | 6.05 |
| P6 | CLB, CBZ, TPM, PHT | 4 | Yes | No | 9 | 0.00 | 1.97 | – | – |
| P7 | LEV, VPA, EsliCBZ | 3 | No | No | 16 | 0.00 | 1.16 | 4.63 | 25.28 |
| P8 | CBZ, PHT, PER, CLB | 4 | Yes | No | 5 | 28.13 | 26.70 | – | – |
| P9 | CBZ, PER, LTG | 3 | No | No | 17 | 2.26 | 44.24 | 0.13 | 1.92 |
| P10 | CBZ, PER | 2 | No | No | 12 | 6.09 | 30.04 | 1.87 | 14.12 |
| P11 | LEV, LTG, CLB, PreGB | 4 | Yes | No | 8 | – | – | 0.00 | 2.05 |
| P12 | LTG, LEV, CBZ | 3 | No | No | 9 | 0.04 | 1.51 | 0.08 | 7.94 |
| P13 | LCS, LEV | 2 | No | No | 16 | 1.09 | 6.88 | 1.82 | 12.01 |
| P14 | LCS, LEV | 2 | No | No | 11 | 8.83 | 25.51 | – | – |
| P15 | TPM, CBZ, LEV | 3 | Yes | No | 11 | 0.06 | 7.20 | – | – |
| P16 | OxCBZ, CBZ, ZNS | 3 | No | No | 11 | – | – | 0.05 | 8.80 |
| P17 | LTG, CBZ, PER | 3 | No | No | 14 | 1.57 | 24.63 | 0.83 | 22.48 |
| P18 | CBZ, LTG, CLB | 3 | Yes | No | 12 | 28.06 | 36.17 | 0.38 | 1.87 |
| P19 | TPM, PER, CBZ | 3 | Yes | No | 12 | – | – | 3.20 | 17.06 |
| P20 | PER, CBZ, ZNS | 3 | No | No | 14 | 0.02 | 3.73 | – | – |

ASM = anti-seizure medication; CBZ = carbamazepine; CLB = clobazam; EsliCBZ = eslicarbazepine acetate; LEV = levetiracetam; LCS = lacosamide; LTG = lamotrigine; PER = perampanel; PHT = phenytoin; PreGB = pregabalin; TPM = topiramate; VPA = valproate; ZNS = zonisamide; OxCBZ = oxcarbazepine. ASMs with known prominent cognitive side effects included topiramate, benzodiazepines and barbiturates. Spike rates are expressed as spikes per minute.
